# Epidemiology of early- and late-onset serious bacterial infections in Australian neonates and infants: A retrospective multicentre study

**DOI:** 10.1101/2024.12.14.24318949

**Authors:** Phoebe CM Williams, Mona Mostaghim, Jackson Harrison, Monica Lahra, Mark Greenhalgh, Matthew O’Sullivan, Michael Maley, Ju-Lee Oei, Archana Koirala, Himanshu Popat, Kei Lui, Brendan McMullan, Pamela Palasanthiran, Alison Kesson, David Isaacs, Adam W Bartlett

## Abstract

**Background:** There has been little decline in neonatal mortality rates over recent decades, and this is now further challenged by the rising prevalence of antimicrobial resistance (AMR). In Australia, the incidence of neonatal sepsis is low on a global scale, yet there are increasingly frequent outbreaks of multidrug-resistant (MDR) infections in neonatal intensive care units, alongside rising rates of colonisation with MDR bacteria.

**Methods:** We analysed positive blood and cerebrospinal fluid (CSF) cultures collected from infants (aged 0 to ≤180 days) across five clinical sites in Australia between 2010 and 2019, to determine evolving antimicrobial susceptibility profiles.

**Results:** After excluding presumed contaminants, we analysed 743 pathogenic bacterial isolates cultured from 624 neonates and infants with early- (≤72 hours), late- (>72 hours to ≤28 days), and very late-onset (>28 days to ≤180 days) infections. *Escherichia coli* (37%) and *Streptococcus agalactiae* (31%) were the primary pathogens responsible for early-onset bloodstream infections, whilst coagulase-negative staphylococci, *E. coli* and *Staphylococcus aureus* were responsible for most infections in older neonates and infants. Antimicrobial susceptibility to currently-recommended empiric regimens remains high; however, gram-negative bacteria – including MDR bacteria – were responsible for an increasing proportion of very late-onset infections over the study period (22% in 2010-2014 versus 34% in 2015-2019; p=0.07).

**Conclusions:** Although empiric antimicrobial regimens remain adequate for most pathogens causing infections in neonates and infants in Australia, there is an increasing burden of invasive infections caused by gram-negative bacteria. Ongoing surveillance is necessary to ensure empiric antimicrobial guidelines remain efficacious and appropriate.

## INTRODUCTION

Over the past three decades there has been global progress in reducing child mortality, yet the decline in neonatal mortality rates has been slower, with nearly half of all childhood deaths now occurring within the neonatal (0-28 day) period.^1^ Serious bacterial infections (often referred to as ‘neonatal sepsis‘) are a leading cause of child mortality, accounting for up to 700,000 newborn deaths each year - the majority of which occur in Southeast Asia and sub-Saharan Africa.^2,3^ However, as antimicrobial resistance (AMR) rises globally, the mortality and disability burden caused by multidrug-resistant organisms (MROs) – which are increasingly responsible for a larger proportion of neonatal infections – requires urgent attention across all healthcare settings.^3^

The newborn period bears the highest lifetime risk of sepsis. ^3^ Multiple risk factors predispose neonates (and infants, particularly those born prematurely) to invasive bacterial infections – including immune system immaturity, poorly developed gastrointestinal and skin mucosal barriers, and exposure to invasive procedures when unwell.^4^ In Australia, neonatal sepsis incidence rates are low on an international scale, yet are gradually increasing;^5^ and the associated morbidity and mortality burden is significant.^6^

Advanced healthcare systems now enable the resuscitation and support of infants from 23-24 weeks’ gestation, and these extremely preterm infants subsequently require a prolonged hospital stay during which they are at high risk of developing late-onset sepsis (with incidence rates up to 15 times that observed term infants).^7,8^ Furthermore, premature infants are frequently prescribed multiple empiric antibiotic courses, reducing their microbiome diversity and increasing the risk of colonisation and infection with MROs.^9^

International observational studies have revealed the epidemiology of neonatal sepsis is rapidly evolving due to the growing global burden of AMR.^10–12^ However, there are scarce published data delineating the epidemiology of neonatal infections in Australian infants, with most published literature focussing on early-onset sepsis (EOS; currently defined as infections occurring at ≤72 hours of age).^6,8^ While EOS is an important cause of neonatal morbidity and mortality, understanding the burden of late-onset sepsis (LOS, occurring at >72h of age) is increasingly important as more premature infants survive and require prolonged hospitalisation.^13^

In Australia and New Zealand, LOS occurs in 44% of neonates born at <24 weeks gestational age who are admitted to tertiary neonatal intensive care units (NICUs), and in 17% of admitted infants born at 26-27 weeks gestational age.^14^ While LOS can occur secondary to perinatally-acquired (vertically-transmitted) pathogens, it is often the consequence of acquisition of MROs via colonisation with bacteria present within the hospital setting.^15^ However, there is increasing evidence to suggest hospital-acquired bacteria may also be responsible for a significant burden of EOS, challenging the traditional rationale behind the empirical antibiotic regimens currently-recommended to treat EOS and LOS, which are based on historical data around causative bacteria.^16^

Australia’s therapeutic guidelines recommend benzylpenicillin plus gentamicin as empiric antibiotics to treat EOS (or benzylpenicillin plus cefotaxime if meningitis is suspected),^17^ and an aminopenicillin plus gentamicin for infants with community-acquired LOS (cefotaxime where meningitis is suspected), with the addition of vancomycin if there is an epidemiological risk factor for methicillin-resistant *Staphylococcus aureus.* For infants aged >2 months with septic shock or meningitis, these guidelines recommend gentamicin, cefotaxime/ceftriaxone and vancomycin for infants presenting from the community.^18^ Most hospital- and state-based guidelines across Australia recommend neonates (and infants hospitalised since birth) with LOS are treated with flucloxacillin and gentamicin, +/- vancomycin.^17,19–21^

Despite strong antimicrobial stewardship programs across Australia’s healthcare settings, there are emerging challenges with both MRO neonatal colonisation and MRO outbreaks in NICUs,^13,22–24^ which are increasingly difficult to treat.^25,26^ As the burden of AMR evolves and neonates are resuscitated at earlier gestations, significant knowledge gaps remain in understanding the epidemiology of serious bacterial infections in neonates and infants – including evaluation as to whether the current empiric antibiotic recommendations remain appropriate.^27,28^ We aimed to address this evidence gap by analysing the contemporary bacterial pathogens (and their evolving antibiotic susceptibility profiles) responsible for both early- and late-onset serious bacterial infections in neonates and infants in New South Wales (NSW), Australia’s most populous state.

## METHODS

We conducted a multicentre retrospective observational study across five clinical settings in NSW, Australia (including one rural hospital, and four tertiary urban hospitals). Laboratory databases were interrogated to systematically evaluate positive blood and cerebrospinal fluid (CSF) cultures collected between 2010-2019 (inclusive) in infants aged ≤180 postnatal days of age. We stratified our analysis of culture-positive infections to (i) the early neonatal period (≤72 hours), (ii) the late neonatal period (>72 hours to ≤28 days), and (iii) very late onset infections (>28 days to ≤180 days). State-wide data were used to ascertain denominator data for the number of livebirths within each of the study hospitals.^20^

Cultures collected within 14 days of an index culture isolating the same pathogen were considered duplicates and removed, alongside a pre-defined list of likely contaminant bacteria (Supplementary Figure 1). Coagulase-negative staphylococci (CoNS) isolates were only retained as bloodstream infection pathogens in infants aged >72 hours of age who were admitted to the neonatal intensive care, or in CSF isolates where review of clinical data confirmed the occurrence of meningitis. CoNS isolates from neonates admitted from the community or in postnatal wards were removed as presumed contaminants.

Antimicrobial susceptibility data were attained following testing performed using automated methods (Vitek2®, BioMérieux or BD Phoenix™, Becton Dickinson, USA) and categorised by CLSI,^29^ EUCAST,^30^ or CDS methodology;^31^ then extracted from laboratory systems and evaluated against a pre-defined list of antibiotics (Supplementary Table 1). Pathogens with (non-inherent) non-susceptibility to three or more classes of antibiotics were classified as multidrug-resistant. Changes in causative bacteria and antimicrobial susceptibility were assessed by comparing the periods between January 2010-December 2014 and January 2015-December 2019.

Data cleaning and statistical analysis were performed in the R programming environment (R Core Team, 2023). Continuous data were reported as median and interquartile ranges, while categorical variables were reported as numbers and percentages, and compared using Chi-squared or Fisher exact tests. Poisson regression was used to assess trends in incidence over time. Ethical approval was obtained via the Sydney Children’s Hospital Network Human Research Ethics Committee (2020/ETH01847).

## Results

We analysed 743 pathogenic bacterial isolates collected from 624 infants across five hospitals during the study period (Figure 1; Table 1; Supplementary Figure 1). This included 464 isolates from neonates and infants admitted to Nepean and Westmead Hospital in urban NSW, and Wagga Wagga Base Hospital in rural NSW, between 2010-2019 (inclusive); and 279 isolates from neonates and infants admitted to The Royal Hospital for Women and Sydney Children’s Hospital in Eastern Sydney from 2015-2019 (a shorter period than the pre-defined study period, due to laboratory information system changes for the pathology service associated with these hospitals).

**Figure 1.**
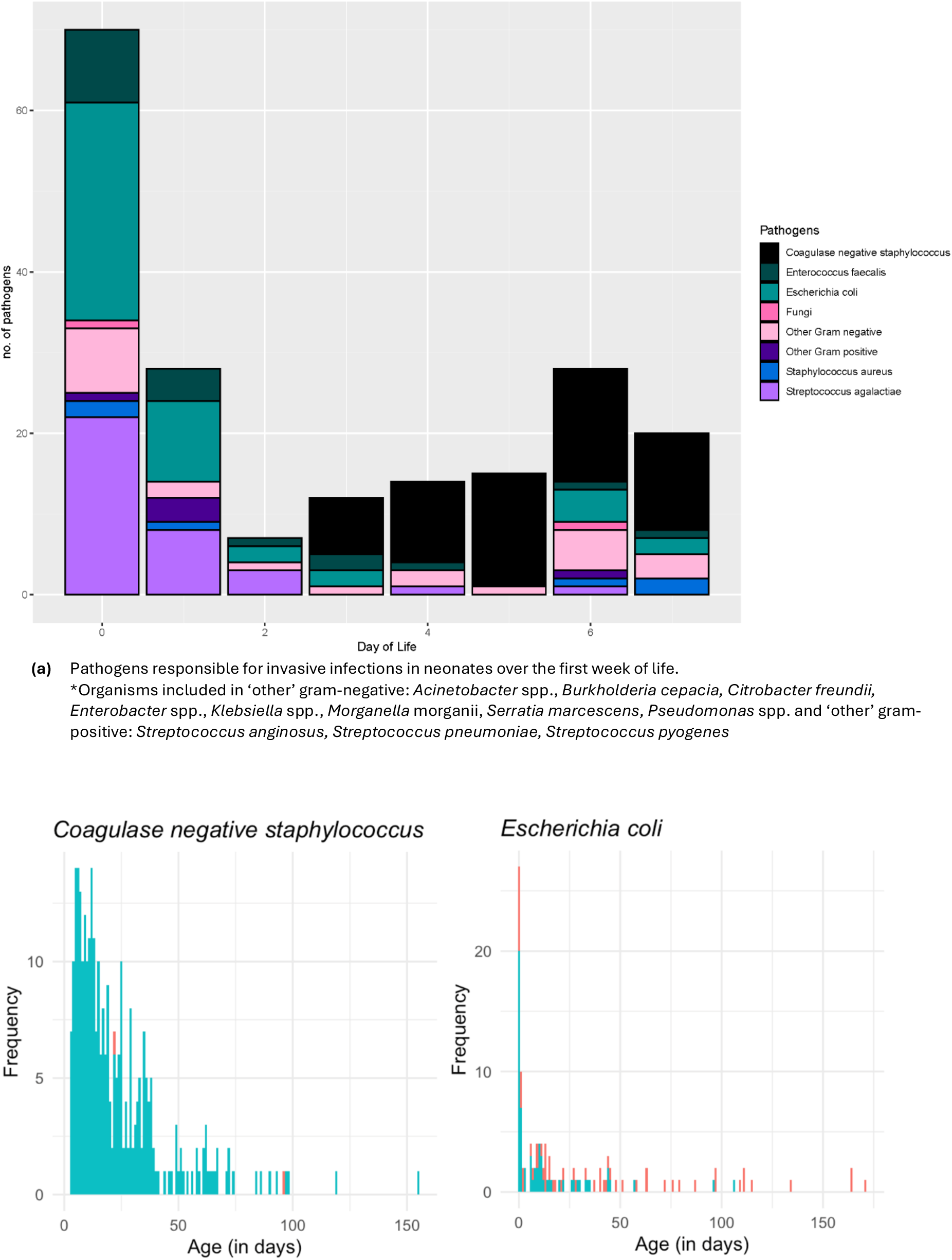
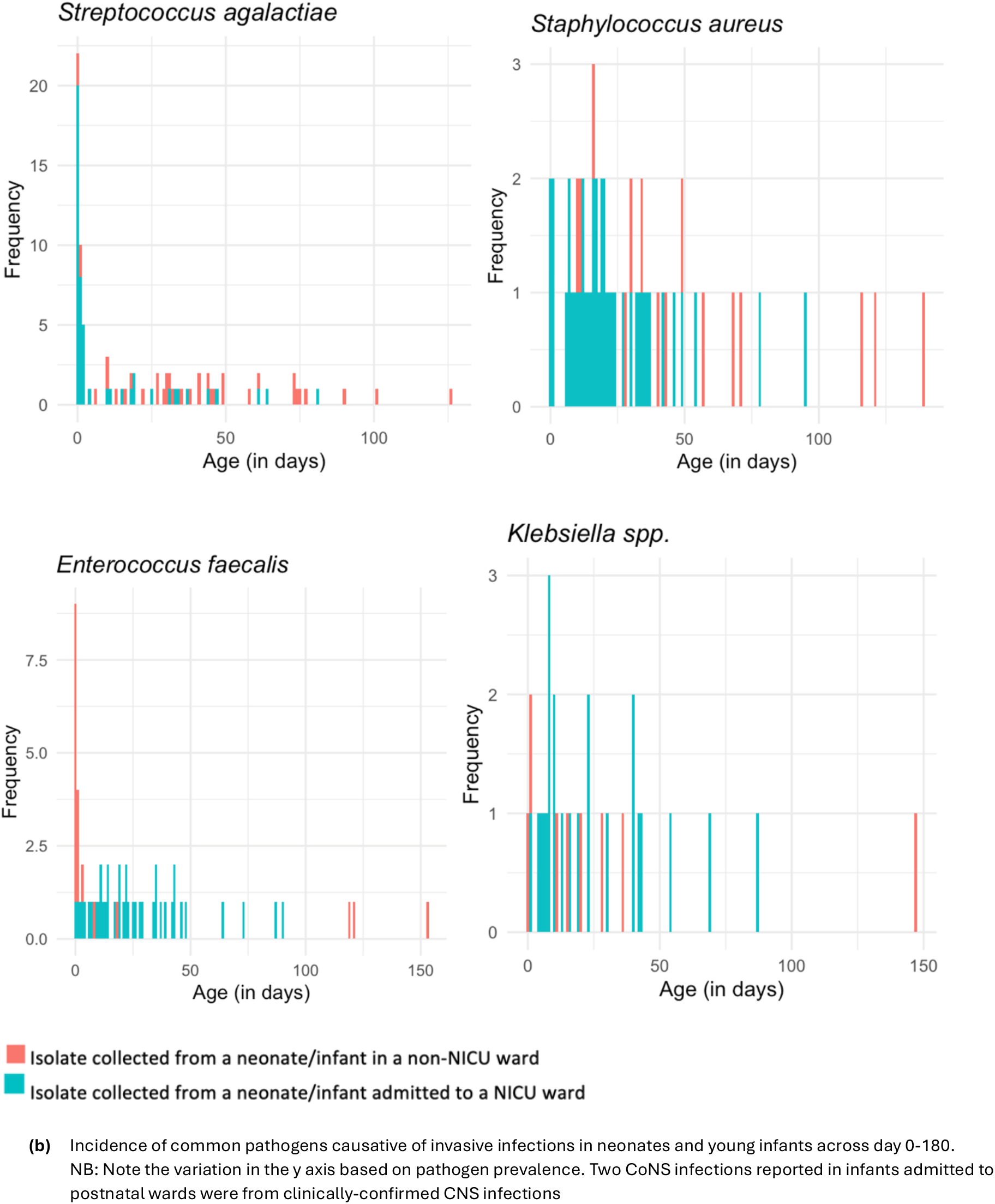
Incidence of common pathogens causing infections in neonates and infants by day of postnatal age; across day 0 to 7 (a); and day 0 to 180d (b)

**Table 1:** Demographic characteristics.

|  | **Total** | **Day 0-2** | **Day 3-27** | **Day 28-180** |
| --- | --- | --- | --- | --- |
|  | n (%) or median (IQR) |  |  |  |
| **Number of infection episodes** | **705** | 106 | 365 | 234 |
| **Patients** | **624** | 103 | 342 | 211 |
| **Gender** |  |  |  |  |
| Female | **291 (41)** | 41 (39) | 152 (42) | 98 (42) |
| Male | **406 (58)** | 57 (54) | 213 (58) | 136 (58) |
| Unknown | **8 (1)** | 8 (7.5) | 0 | 0 |
| **Age** | **16 (7-35)** | 0 (0-1) | 12 (8-18) | 45 (35-70.5) |
| **NICU** | **543 (77)** | 75 (71) | 321 (88) | 147 (63) |
| **Year** |  |  |  |  |
| 2010-2014 | **252 (36)** | 38 (36) | 140 (38) | 74 (32) |
| 2015 - 2019 | **453 (64)** | 68 (64) | 225 (62) | 160 (68) |
| **Specimen type** |  |  |  |  |
| Bloodstream | **669 (95)** | 101 (95) | 353 (97) | 215 (92) |
| Cerebrospinal Fluid | **36 (5)** | 5 (5) | 12 (3) | 19 (8) |
| **Polymicrobial Infections<sup>^</sup>** |  |  |  |  |
| Bloodstream | **34 (5)** | 4 (4) | 17(5) | 13 (6) |
| Cerebrospinal Fluid | **1 (<1)** | 0 | 0 | 1 (5) |
| **Hospital** |  |  |  |  |
| Nepean Hospital | **137 (19)** | 30 (28) | 70 (19) | 37 (16) |
| Royal Hospital for Women | **165 (23)** | 17 (16) | 96 (26) | 52 (22) |
| Sydney Children's Hospital | **98 (14)** | 8 (8) | 27 (7) | 63 (27) |
| Wagga Wagga Base Hospital | **18 (3)** | 8 (8) | 7 (2) | 8 (3) |
| The Children's Hospital at Westmead | **287 (41)** | 48 (45) | 165 (45) | 74 (32) |

<sup>^</sup> % of specimen type

Bloodstream infections were responsible for most infection episodes (95%, 669/743), with 35 CSF infections also identified (of 743, 5%). There were 34 polymicrobial bloodstream infections (≥ two pathogenic bacteria isolated), and one CSF specimen yielded three pathogenic bacteria (Table 1). Most isolates were obtained from neonates or infants admitted to NICUs (73%, 543/743). The incidence of EOS across the study period varied from 0.6 per 1,000 livebirths to 1 per 1,000 livebirths (Supplementary Table 2), with no significant trend in EOS incidence rates observed across the study period (0.05% change in incidence per year, 95%CI -7.1% to 8.9%).

### Early onset neonatal infections (≤72 hours of age)

Fifteen percent (106/705) of infections occurred in the EOS period (≤72 hours of age), with a median age of infection onset in this cohort of 0 days (IQR 0-1 day). Bloodstream infections were caused by approximately equal proportions of gram-negative (48%) and gram-positive (51%) bacteria, and there was one episode of fungaemia (due to *Candida albicans)* (Table 2; Figure 1).

**Table 2:** Bacteria causative of bloodstream and CSF infections in neonates and infants 0-180 days of age. Data are n (%); ^Coagulase negative staphylococcus excluded when reported on general wards or day 0-2 of life

|  | Blood |  |  | Cerebrospinal fluid |  |  | Total |
| --- | --- | --- | --- | --- | --- | --- | --- |
|  | Day 0-2<br>n=105 | Day 3-27<br>n= 370 | Day 28-180<br>n= 230 | Day 0-2<br>n= 5 | Day 3-27<br>n= 12 | Day 28-180<br>n= 21 |  |
| **Gram-negative bacteria** | **50 (48)** | **91(25)** | **68 (30)** | **0 (n/a)** | **5 (42)** | **11 (52)** | **225** |
| *Escherichia coli* | 39 (37) | 39 (10) | 35 (15) | 0 | 3 (25) | 4 (19) | 120 |
| *Klebsiella* spp. | 3 (3) | 17 (5) | 10 (4) | 0 | 0 | 1 (5) | 31 |
| *Pseudomonas* spp. | 2 (2) | 10 (3) | 4 (2) | 0 | 1 (8) | 2 (9.5) | 19 |
| *Enterobacter* spp. | 1 (1) | 14 (4) | 8 (3) | 0 | 1 (8) | 1 (5) | 25 |
| *Serratia marcescens* | 1 (1) | 3 (1) | 4 (2) | 0 | 0 | 1 (5) | 9 |
| *Acinetobacter* spp. | 1 (1) | 6 (2) | 4 (2) | 0 | 0 | 0 | 11 |
| Other gram-negative bacteria | 3 (3) | 2 (0.5) | 3 (1) | 0 | 0 | 2 (10) | 10 |
| **Gram-positive bacteria** | **54 (51)** | **278 (75)** | **162 (70)** | **5 (100)** | **7 (58)** | **10(48)** | **516** |
| Coagulase negative staphylococcus^ | n/a | 209 (56) | 88 (38) | 0 | 3 (25) | 2 (10) | 302 |
| *Enterococcus faecalis* | 14(13) | 23 (6) | 19 (8) | 0 | 1 (8) | n/a | 57 |
| *Streptococcus agalactiae* | 33(31) | 15 (4) | 28 (12) | 4(80) | 2 (17) | 5 (24) | 87 |
| *Staphylococcus aureus* | 3 (3) | 28 (8) | 22 (10) | 1(20) | 1 (8) | 3 (14) | 58 |
| Other gram-positive bacteria | 4 (4) | 3 (1) | 5 (2) | 0 | 0 | 0 | 12 |
| **Fungal** | **1 (1)** | **1 (<1)** | **0** | **0** | **0** | **0** | **2** |
| *Candida albicans* | 1(1) | 1 (<1) | 0 | 0 | 0 | 0 | 2 |

*Escherichia coli* (37%), *Streptococcus agalactiae* (31%) and *Enterococcus faecalis* (13%) were identified as the bacteria most frequently responsible for EOS (Figure 1). The proportion of gram-negative bacteria responsible for EOS was similar across the study period (58% over 2010-2014 versus 43% over 2015-2019, p=0.13). Neonates who required admission to the NICU for higher-level care were more likely to have *S. agalactiae* infections (41%, versus 12% of infants admitted to postnatal wards), while neonates with *E. faecalis* infections were more likely to be admitted to the postnatal ward (32% versus 4% of infants admitted to the NICU). All CSF infections (n=5) in this age group were caused by gram-positive bacteria (*S. agalactiae* and *S. aureus*) (Table 2).

### Late onset neonatal infections (day 3 to <28 days)

Most of the culture-positive infections identified in our study (52%, 365/705) occurred in the late-onset neonatal period (day 3 to <28 days), at a median age of 12 days (IQR 8 to 18). Within this cohort, there were 353 episodes of bacteraemia (incorporating 370 isolates, including 17 polymicrobial infections) and 12 CSF infections.

Gram-positive bacteria predominated as causative of bloodstream infections (n=278, 75%), largely due to a significant burden of CoNS isolated from neonates admitted to the NICU (209/370, 56%), followed by *S. aureus* (8%), *E. faecalis* (6%) and *S. agalactiae* (4%) infections. One episode of fungaemia was reported (due to *C*. *albicans).* The most frequently isolated gram-negative bacteria causative of late-onset neonatal bloodstream infections was *E. coli* (10%, 39/370), followed by *Klebsiella* spp. (5%, 17/370) and *Enterobacter cloacae* complex (14/370, 4%). The proportion of bloodstream infections caused by gram-negative bacteria was largely consistent across the two periods (21% in 2010-2014 and 27% in 2015-2019, p = 0.26). The 12 CSF infections were caused by *E. coli, S. agalactiae, S. aureus, Pseudomonas. aeruginosa, Enterobacter* spp. and CoNS (Table 2).

### Very late onset infections in infants (day 28-180 inclusive)

One-third of isolates included in our analysis (234/705, 33%) were cultured from infants aged 28-180 days (median age of infection at 45 days, IQR 35 to 71). Gram-positive bacteria predominated as causative of bloodstream infections in this age group (162/230, 70%), again due to a significant burden of CoNS bloodstream infections in neonates admitted to the NICU (38%, 88/230), alongside very late-onset *S. agalactiae* infections (12%, 28/230), and bloodstream infections caused by *S. aureus* (10% 22/230) and *E. faecalis* (8%, 19/230).

Among gram-negative bacteria, *E. coli* was responsible for 15% of all bloodstream infections (35/230) in this cohort, followed by *Klebsiella* spp. (10/230, 4%) and *Enterobacter* spp. (8/230, 3%). Across all hospitals, an increasing proportion of late-onset bloodstream infections were caused by gram-negative bacteria over the study period (22% over 2010-2014 versus 34% over 2015-2019; p=0.07). Among the pathogens causative of the 21 CSF infections in this cohort, *S. agalactiae* (5/21) predominated, followed by *E coli* (4/21, 19%) and *S. aureus* (3/21).

### Antimicrobial Susceptibility Profiles

#### i. Gram-positive bloodstream infections

*S. agalactiae* was a prominent cause of bloodstream infections across the study population (n=76 isolates), with all tested isolates susceptible to penicillins (n=76) and third-generation cephalosporins (cefotaxime/ceftriaxone, n=11/11 tested; Supplementary Table 4). Most *S. agalactiae* isolates were also susceptible to clindamycin (27/39, 70%) and erythromycin (32/44, 73%). Most *S. aureus* isolates were methicillin-susceptible (44/53, 83%), and over 90% were susceptible to clindamycin (42/46) or trimethoprim-sulfamethoxazole (25/27); whilst all tested *S. aureus* isolates were susceptible to vancomycin (40/40). There were no statistically significant susceptibility changes among *S. aureus* isolates across the study period (Supplementary Table 3). Of 56 *E. faecalis* isolates with susceptibility data available, all tested against ampicillin (39/39) and vancomycin (55/55) were susceptible.

#### i. Gram-negative bloodstream infections (BSIs)

*E. coli* was the primary cause of gram-negative bloodstream infections in our study population (n=113; Figure 2). Of those tested, 92% demonstrated susceptibility to third-generation cephalosporins (ceftriaxone/cefotaxime; 100/109), 89% were susceptible to gentamicin/tobramycin (101/113), and 45% were susceptible to aminopenicillins (ampicillin/amoxicillin, 33/74). All *E. coli* BSI isolates tested were susceptible to amikacin (75/75) and meropenem (78/78). Trimethoprim-sulfamethoxazole susceptibility decreased for *E. coli* isolates over the study period (from 85% [33/39] in 2010-2014 to 65% [22/34] in 2015-2019, p=0.05), yet no other statistically significant changes in AST results were observed.

**Figure 2.**
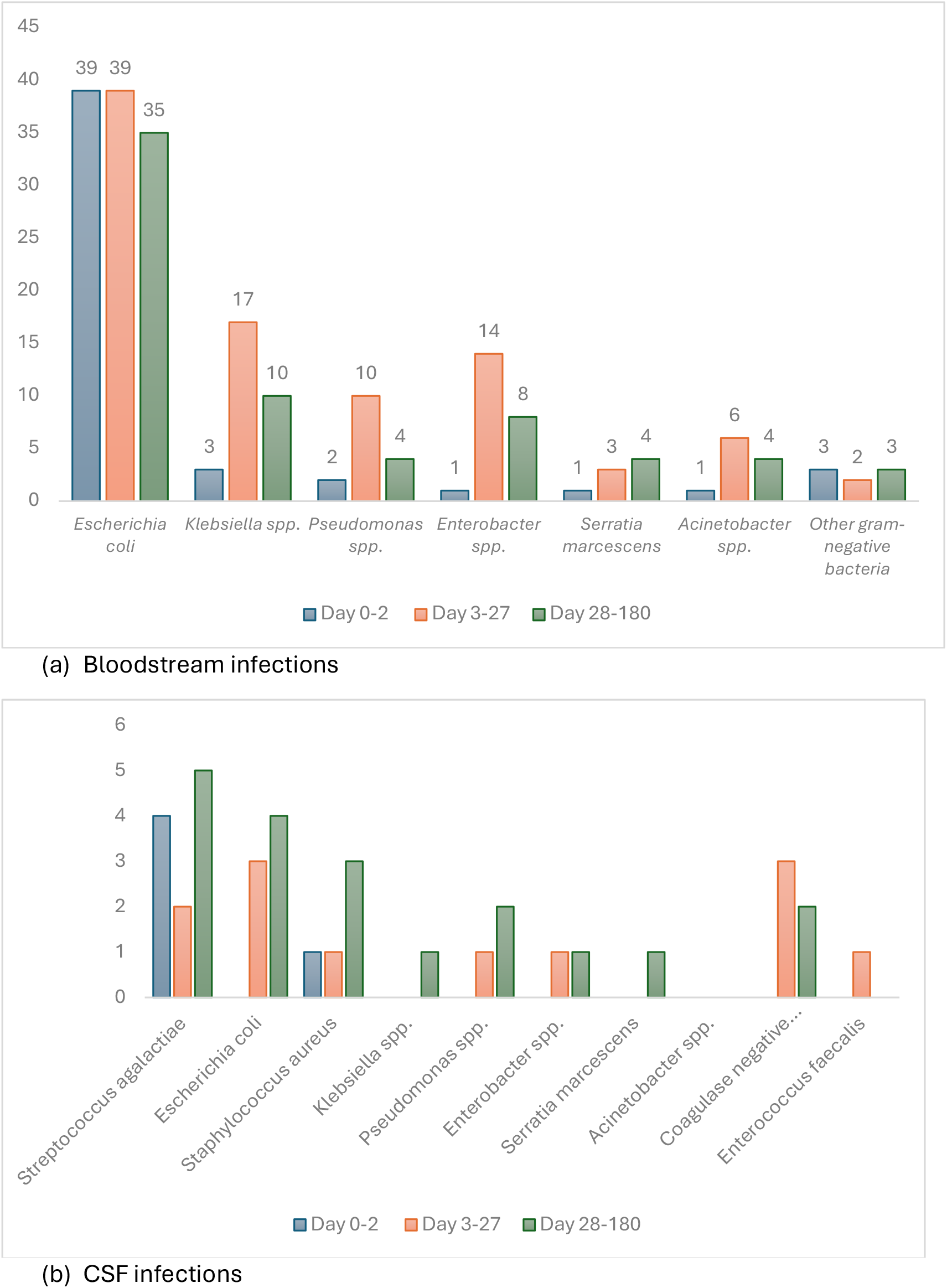
Bacterial pathogens causative of (a) bloodstream (b) CSF infections in neonates and infants.

There were 14 episodes of MDR *E. coli* bacteraemia (defined as acquired non-susceptibility to at least one antibiotic across three or more antimicrobial classes), including eight (57%) which were extended-spectrum beta-lactamase producing pathogens). Most of these infections (9/14) occurred at the two hospitals in Western Sydney as sporadic infections (not associated with outbreaks) in the latter part of the study period (2015-2019, versus 5/14 in 2010-2014, p=0.18)

*Enterobacter cloacae* complex was the second most common gram-negative bacteria responsible for bloodstream infections (n=23). Most tested isolates were susceptible to meropenem (95%, 19/20), cefepime (14/16, 88%), gentamicin/tobramycin (87%), amikacin (16/16, 100%), ciprofloxacin (19/19) and trimethoprim-sulfamethoxazole (17/17). There were two MDR *E. cloacae* bloodstream infections (including one carbapenem-resistant infection), which occurred in 2017 and 2019 in infants admitted to the hospitals in Central Sydney. All tested *Klebsiella* spp. and *Pseudomonas aeruginosa* isolates had high rates of susceptibility to all antibiotics assessed (Supplementary Table 4).

#### iii. Antimicrobial susceptibility in CSF infections

Gram-negative CSF infections were primarily caused by *E. coli* (7/16. 44%), with all tested CSF isolates (n=7) universally susceptible to third-generation cephalosporins (ceftriaxone/cefotaxime) and aminoglycosides (Supplementary Table 5). *Enterobacter cloacae* complex CSF isolates (n=2) and *Pseudomonas aeruginosa* CSF isolates (n=2) were susceptible to aminoglycosides (gentamicin/tobramycin) and later-generation cephalosporins. Gram-positive CSF infections were primarily caused by *S. agalactiae*, with all isolates susceptible to penicillin (11/11) and ceftriaxone (5/5), as anticipated. One *S. aureus* CSF isolate was methicillin-resistant (of 5, 20%; Supplementary Table 6).

#### iv. Antimicrobial susceptibility to commonly-prescribed antibiotics

Table 3 summarises the susceptibility of the most frequently isolated pathogens to commonly-prescribed empiric antimicrobials. *E coli* isolates from infants with very late-onset infections were less likely to be susceptible to ampicillin than those attained from neonates, though gentamicin and third-generation susceptibility remained high across all age cohorts. Methicillin-resistant *S. aureus* evolved with increasing postnatal age, with no tested isolates exhibiting methicillin-resistance in the EOS period, while 34% (day 3-27) and 13% (day 28-180) of isolates revealed methicillin-resistance in older neonates and infants (respectively). *E. faecalis* isolates were all susceptible to vancomycin for the 0-2 day cohort, yet declined to 83% in older neonates (3-27 days) and 89% in infants (28-180 days)

**Table 3:**
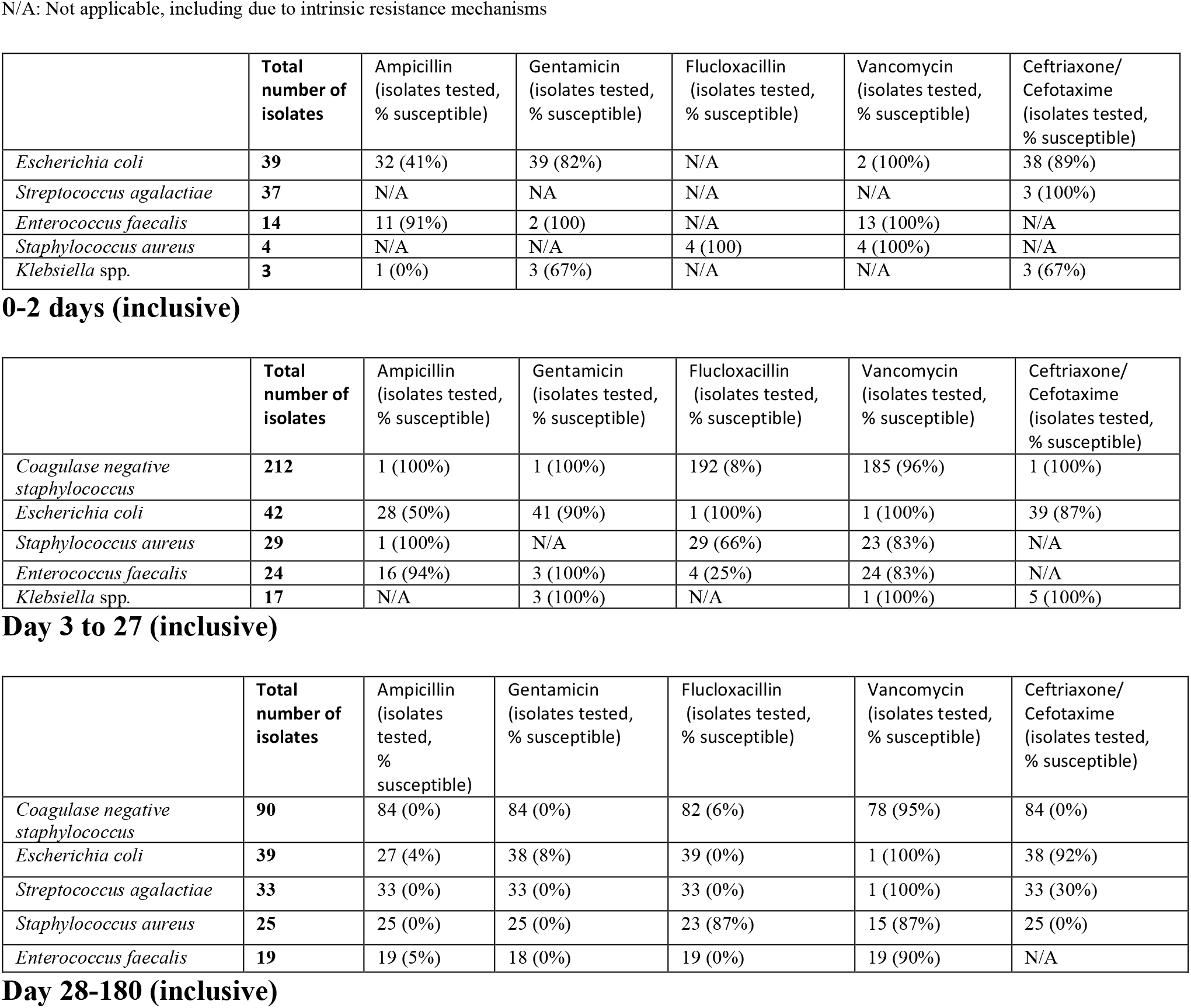
Susceptibility profiles of the five most common pathogens causative of invasive (bloodstream and CSF) infections Summarised according to commonly-recommended antimicrobial combinations, by age cohort. N/A: Not applicable, including due to intrinsic resistance mechanisms

## DISCUSSION

Our multicentre study evaluated the pathogens responsible for serious bacterial infections in neonates and infants (up to 6 months’ postnatal age) across five hospitals in urban and rural NSW, Australia. Our study revealed a significant burden of serious bacterial infections occur beyond the neonatal period, and that many of these infections are due to gram-negative bacteria, with a rising prevalence of multidrug-resistance evident over time. As healthcare settings are increasingly able to resuscitate and support extremely premature infants, prolonged hospital stay beyond the neonatal period necessitates the close monitoring of the epidemiology of these infections, particularly as the burden of AMR rises globally.

Our analysis is unique by way of inclusion of serious infections occurring beyond the neonatal period. Most published data in this field focuses on the epidemiology of early-onset neonatal infections, yet our study has revealed a substantial burden of serious bacterial infections occurring in infants. Many of these infants are prematurely-born and vulnerable to hospital-acquired infections, emphasising the need for ongoing surveillance and evaluation of the appropriateness of empiric antibiotic regimens, to ensure they remain targeted to the most likely causative pathogens. This is crucial in the context of the known challenges in confirming microbiological diagnoses in neonates and infants with invasive infections, which often limits the capacity for directed therapy.^32^

Our data reveal adequate levels of susceptibility to currently-recommended and commonly-prescribed empiric antimicrobial combinations, suggesting local empiric antibiotic guidelines for sepsis and meningitis remain appropriate, in alignment with other observational studies conducted in high-income settings.^33,34^ However, MDR gram-negative infections were more prevalent in the latter part of our study period, and invasive bacterial infections occurring beyond the EOS period were more likely to exhibit antibiotic resistance. Furthermore, more than 10% of *E. coli* isolates in our study were non-susceptible to gentamicin, which has prompted some NICUs in Australia to alter their empirical aminoglycoside to amikacin over recent years.

Our study revealed interesting findings regarding the bacteria responsible for serious bacterial infections in neonates and infants admitted to various ward settings, with those admitted to higher-dependency settings (ie, NICUs) more likely to have infections with gram-negative pathogens (*E coli*), while infants admitted to paediatric or postnatal wards at the time of their culture collection were more likely to have gram-positive infections (*Enterococcus* spp.). Whether this reflects differences in clinical acuity associated with pathogenic bacterial virulence is uncertain, and exemplifies the need for detailed prospective observational studies that incorporate both microbiological *and* clinical data.

Late-onset bloodstream infections accounted for over half of all infections in our cohort, with a substantial proportion caused by CoNS in older neonates and infants admitted to NICUs. This finding aligns with existing literature, highlighting the significant role of CoNS in NICU-acquired infections, particularly in premature infants requiring long lines and prolonged hospitalisation.^35^ However, there was also a notable burden of gram-negative bacteria causative of late-onset infections in our cohort, highlighting the ongoing need for vigilant infection prevention and control (IPC) measures in NICU settings, to mitigate nosocomial transmission of these bacteria given their high propensity to acquire resistance mechanisms.^35^ Although MDR infections were relatively infrequent in our cohort, their presence emphasises the critical importance of ongoing IPC and antimicrobial stewardship efforts in NICU settings. On a promising note, our study revealed a very low burden of invasive fungal infections in neonates and infants, affirming the importance (and success) of fungal prophylaxis programs across NICU settings in Australia.^36^

One of the main limitations of our study is the lack of clinical outcome data, which is a common challenge noted in AMR surveillance literature, and highlights the need for future prospective surveillance studies that capture both microbiological and clinical data. Another limitation in our project is the change in laboratory information systems that occurred across two hospital sites during our study period, limiting the availability of data to a shorter timeframe for those these sites. Challenges in integrating laboratory information systems to easily procure data to examine epidemiological trends in infections is a known issue in the published literature; and strategies to address this have been developed – including for resource-constrained healthcare settings, where the burden of neonatal infection is greatest.^37^

Despite these limitations, to our knowledge this study is the most comprehensive analysis of the pathogens responsible for neonatal and infant bloodstream and CSF infections in Australia. We have revealed a low yet notable burden of serious bacterial infections caused by antibiotic-resistant bacteria, which will require close monitoring - particularly in the context of the very high rates of MDR gram-negative bacteria causing a significant neonatal sepsis burden in Australia’s neighbouring Southeast Asia region.^10–12,38,39^ Furthermore, our analysis of the pathogens causing serious bacterial infections by day of life underscores the importance of continued surveillance of pathogens presumed to be responsible for EOS, both in Australia and globally.^40^

Our study revealed a stable incidence of EOS, with *E coli* and *S agalactiae* identified as the predominant causative agents of EOS. While this is consistent with international data from other high-income healthcare settings, it lies in stark contrast to the bacteria causative of EOS in resource-strained healthcare settings, where MDR gram-negative infections predominate.^10–12^ Ongoing surveillance of the epidemiology of neonatal infections is clearly necessary as more babies are born and survive prematurely, and this data should incorporate wards across all geographic regions. Close monitoring of evolving susceptibility profiles in invasive infections as the burden of AMR increases is vital to improving neonatal health outcomes, and should include both microbiological and clinical data, to evaluate the impact of MDR infections on clinical outcomes.

Alongside improved clinical and microbiological surveillance, further research that identifies and evaluates effective and targeted IPC interventions to reduce late-onset infections in neonates is also necessary, to reduce the burden of nosocomial infections. Furthermore, evaluation of the transmission dynamics of MDR pathogens within NICUs, and how colonisation and invasive infection may interact, is needed to ensure any IPC interventions can be targeted and effective. Finally, interventional studies to identify novel strategies to reduce the morbidity and mortality of neonatal infections (such as the role of steroids in neonatal meningitis, or the use of probiotics to protect the infant microbiome) are necessary, concurrent to an enhanced focus on ensuring neonates can access novel antibiotics needed to effectively treat MDR infections. Together, these strategies may reduce the currently unacceptable mortality burden caused by neonatal sepsis globally.^41^

## Data Availability

All data produced in the present study are available upon reasonable request to the authors.

## Supplementary Data

**Supplementary Figure 1.**
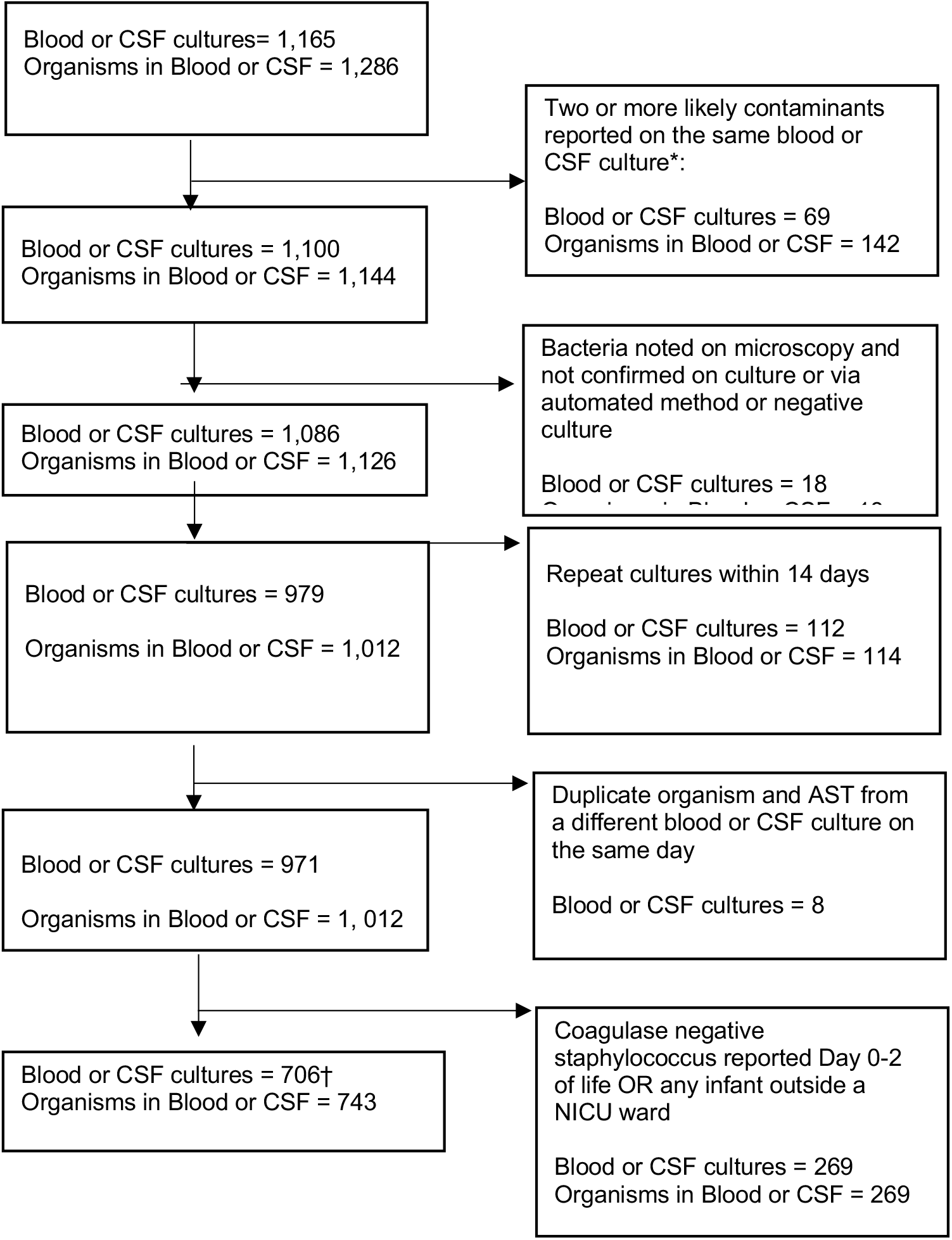
Flow chart of isolates included in the analysis * Contaminants excluded: *Actinomyces naeslundii, Actinomyces odontolyticus, Aerococcus viridans, Bacillus cereus, Bacillus* spp. *Bacteroides vulgatus,* Coagulase negative staphylococcus (CoNS; in infants not admitted to PICU or <72h) *, Corynebacterium minutissimum, Corynebacterium striatum, Empedobacter brevis, Enterococcus durans, Enterococcus hirae, Kocuria varians, Lactococcus lactis subspecies lactis, Leuconostoc pseudomesenteroides, Micrococcus luteus, Micrococcus* spp.*, Moraxella osloensis, Morganella morganii, Neisseria cinerea, Paenibacillus* spp*, Paenibacillus urinalis, Raoultella ornithinolytica, Staphylococcus auricularis, Staphylococcus capitis, Staphylococcus caprae, Staphylococcus epidermidis, Staphylococcus haemolyticus, Staphylococcus hominis, Staphylococcus lugdunensis, Staphylococcus saprophyticus, Staphylococcus spp., Staphylococcus warneri, Staphylococcus xylosus, Streptococcus anginosus, Streptococcus constellatus subspecies pharynges, Streptococcus milleri group, Streptococcus mitis/ Streptococcus oralis* group*, Streptococcus parasanguinis, Streptococcus peroris, Streptococcus salivarius, Streptococcus sanguinis, Streptococcus vestibularis, Viridans group Streptococcus.* ^Further likely contaminants removed (CoNS removed as indicated in the flowchart):*Actinomyces naeslundii, Bacillus cereus, Bacillus species, Corynebacterium minutissimum, Corynebacterium striatum, Empedobacter brevis, Enterococcus durans, Enterococcus hirae, Kocuria varians, Lactococcus lactis subspecies lactis, Leuconostoc pseudomesenteroides, Micrococcus luteus, Micrococcus* spp.*, Paenibacillus* spp., *Paenibacillus urinalis, Raoultella ornithinolytica, Streptococcus anginosus, Streptococcus milleri* group*, Streptococcus mitis/ Streptococcus oralis* group*, Streptococcus parasanguinis, Streptococcus peroris, Streptococcus salivarius, Streptococcus sanguinis, Streptococcus vestibularis, Viridans group Streptococcus.* † One duplicate (same day) blood culture result retained for additional antimicrobial susceptibility results only.

**Supplementary Table 1:** Pre-defined bacteria-antibiotic susceptibility profiles evaluated.

|  |
| --- |
| Gram-positive organisms |
| ***Staphylococcus aureus*** |
| flu/dicloxacillin |
| benzylpenicillin |
| clindamycin |
| cotrimoxazole |
| vancomycin |
| **Streptococcus Group A, B, C and G** |
| benzylpenicillin |
| ceftriaxone/cefotaxime |
| clindamycin |
| erythromycin |
| ***Streptococcus pneumoniae*** |
| benzylpenicillin: blood culture: S / I or DS / R |
| benzylpenicillin: CSF culture: S / I or DS / R |
| clindamycin |
| co-trimoxazole |
| ceftriaxone/cefotaxime: Blood culture: S / I or DS / R |
| ceftriaxone/cefotaxime: CSF culture: S / I or DS / R |
| vancomycin |
| **Coagulase negative *Staphylococcus* spp.** |
| flu/dicloxacillin |
| benzylpenicillin |
| clindamycin |
| vancomycin |
| **Other Gram-positive organisms not otherwise specified** |
| benzylpenicillin |
| aminopenicillin |
| ceftriaxone/cefotaxime |
| meropenem |
| vancomycin |
| clindamycin |
| co-trimoxazole |
| Gram-negative pathogens |
| ***Escherichia coli*** |
| aminopenicillin |
| amoxicillin-clavulanate |
| cephalexin |
| ceftriaxone |
| cefotaxime |
| ceftazidime |
| cefepime |
| meropenem |
| co-trimoxazole |
| gentamicin |
| amikacin |
| tigecycline |
| fosfomycin |
| ciprofloxacin |
| ***Klebsiella pneumoniae*** |
| ceftriaxone |
| cefotaxime |
| cefepime |
| ceftazidime |
| ciprofloxacin |
| gentamicin |
| amikacin |
| co-trimoxazole |
| fosfomycin |
| meropenem |
| norfloxacin |
| tigecycline |
| amoxicillin-clavulanate |
| ***Pseudomonas aeruginosa*** |
| amikacin |
| gentamicin |
| meropenem |
| fosfomycin |
| piperacillin-tazobactam |
| ciprofloxacin |
| ceftazidime |
| cefepime |
| ***Acinetobacter spp.*** |
| gentamicin |
| amikacin |
| meropenem |
| tigecycline |
| minocycline |
| cefepime |
| ciprofloxacin |
| co-trimoxazole |
| piperacillin-tazobactam |
| **Other Gram-negative pathogens not otherwise listed** |
| aminopenicillin |
| ampicillin-clavulanate |
| cephalexin |
| ceftriaxone/cefotaxime |
| ceftazidime |
| cefepime |
| meropenem |
| co-trimoxazole |
| trimethoprim |
| gentamicin |
| amikacin |
| tigecycline |
| fosfomycin |
| ciprofloxacin |
| piperacillin-tazobactam |

**Supplementary Table 2:** Early-onset neonatal sepsis incidence rates across the study period for the five hospitals included in the study.

| Year | Total births in study hospitals | Total EOS cases | EOS incidence per 1,000 livebirths |
| --- | --- | --- | --- |
| 2010 | 9152 | 6 | 0.66 |
| 2011 | NA | 6 | NA |
| 2012 | 9683 | 6 | 0.62 |
| 2013 | 9717 | 9 | 0.93 |
| 2014 | 10106 | 8 | 0.79 |
| 2015 | 14211 | 14 | 0.99 |
| 2016 | 14752 | 11 | 0.75 |
| 2017 | 14616 | 13 | 0.89 |
| 2018 | 14200 | 9 | 0.63 |
| 2019 | 14369 | 11 | 0.77 |

NA: Data not available.

**Supplementary Table 3.**
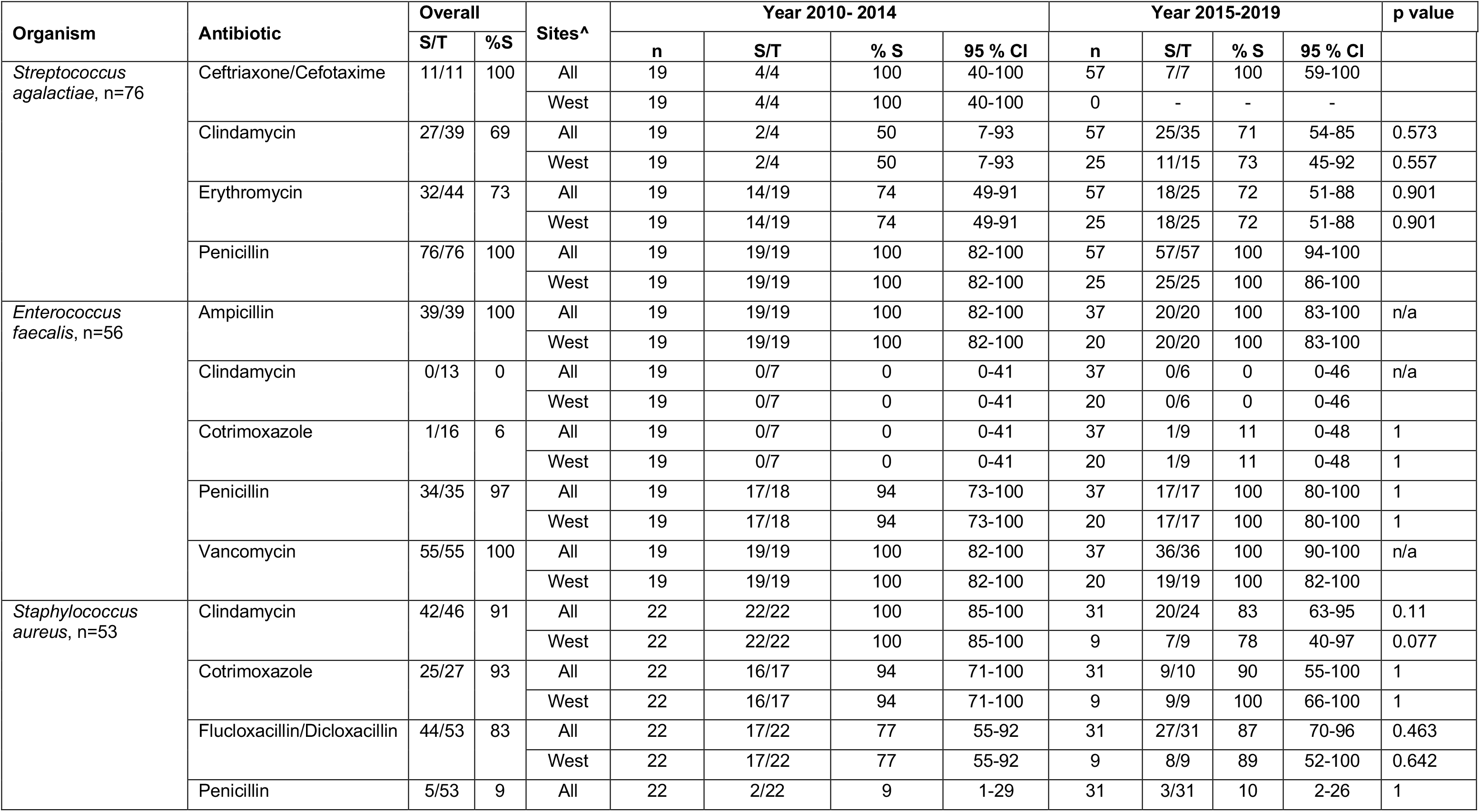
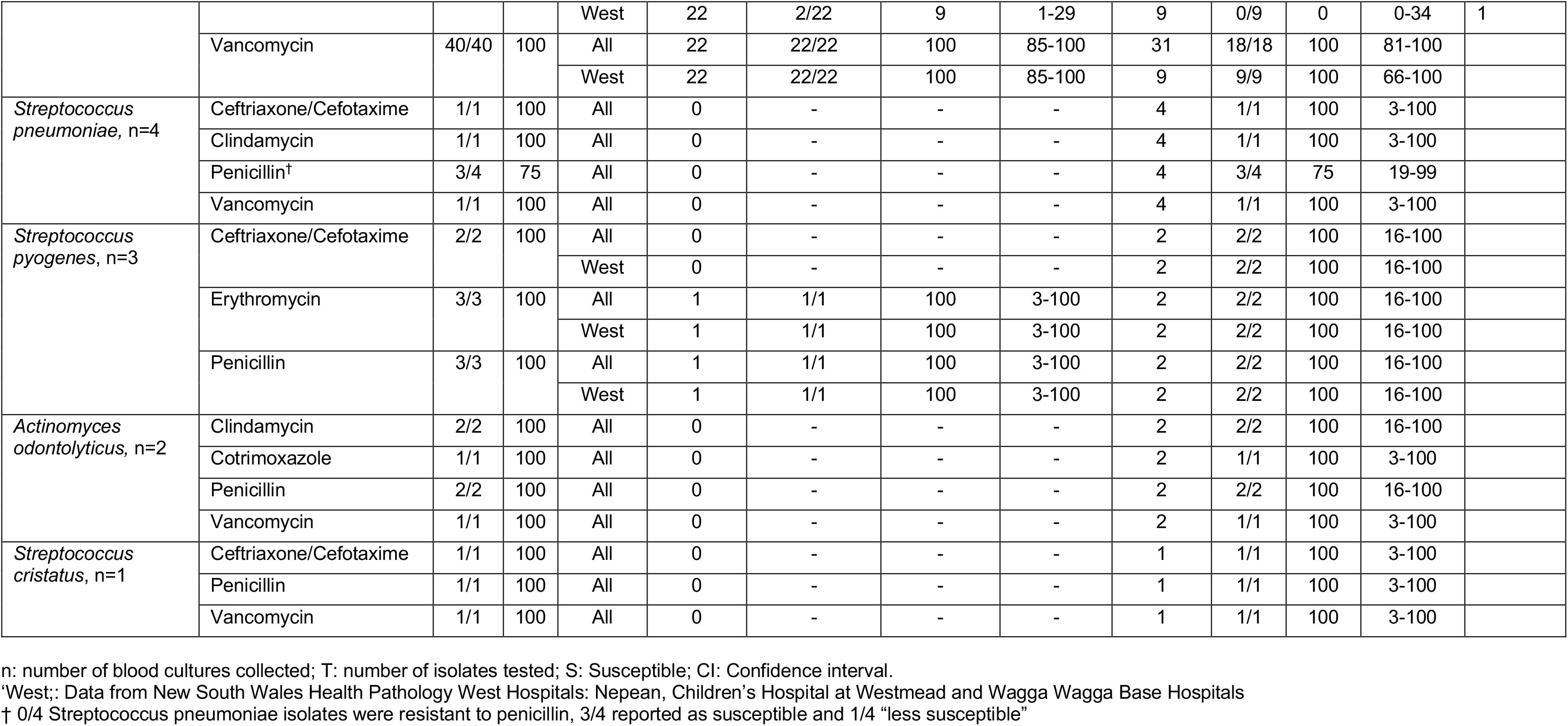
Gram-positive Blood Culture Isolate Antimicrobial Susceptibility Profiles.

**Supplementary Table 4.**
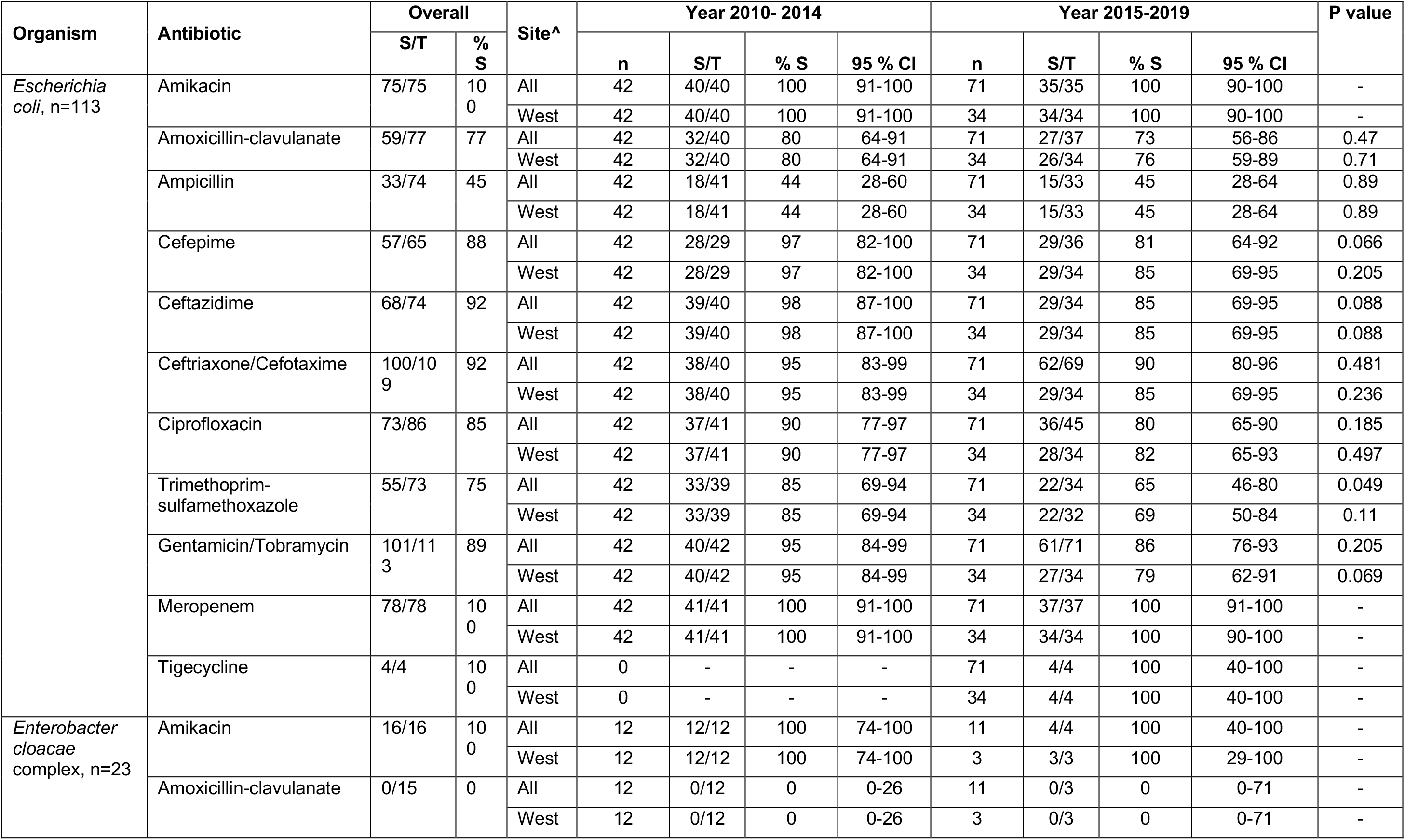
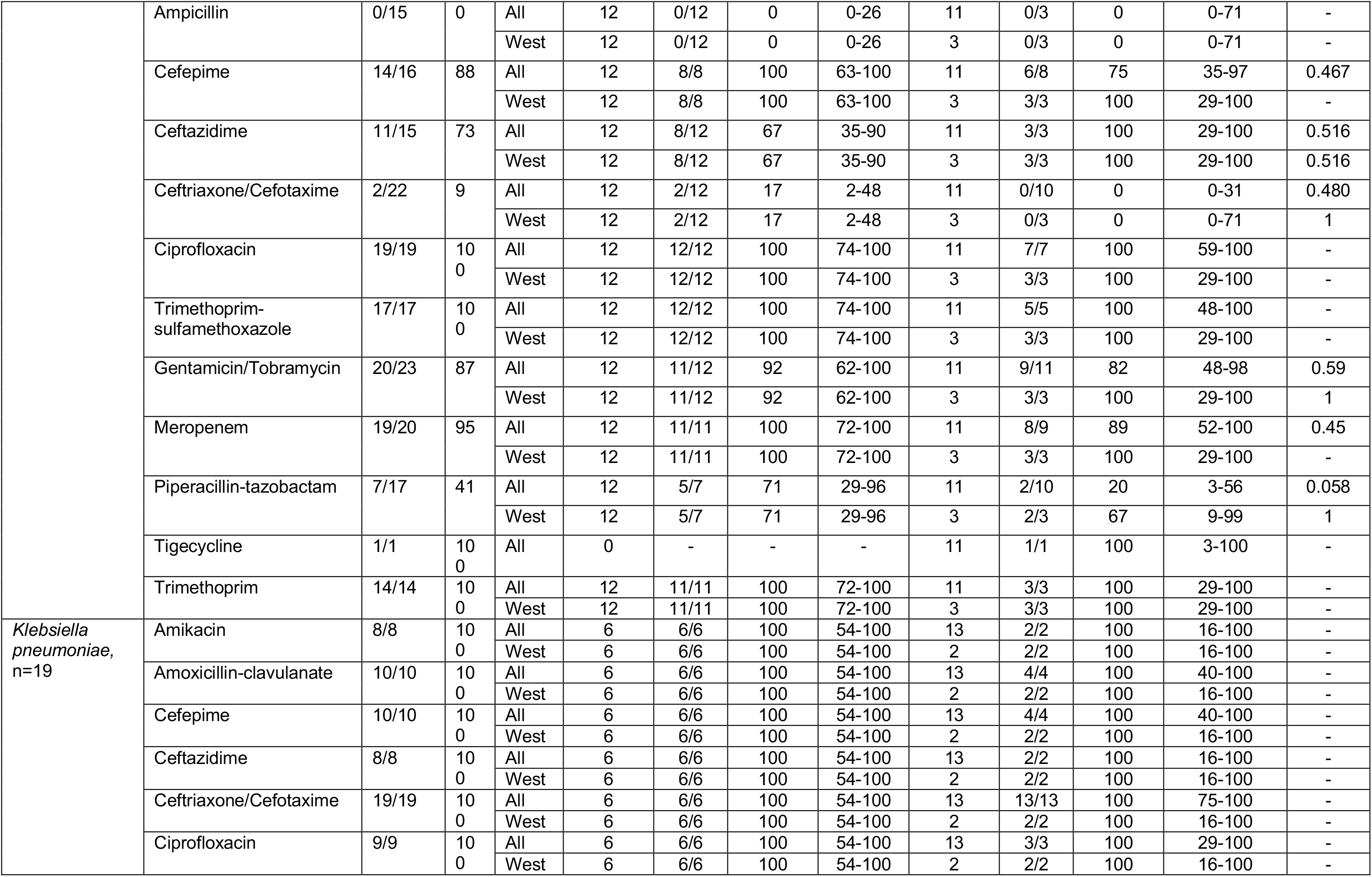
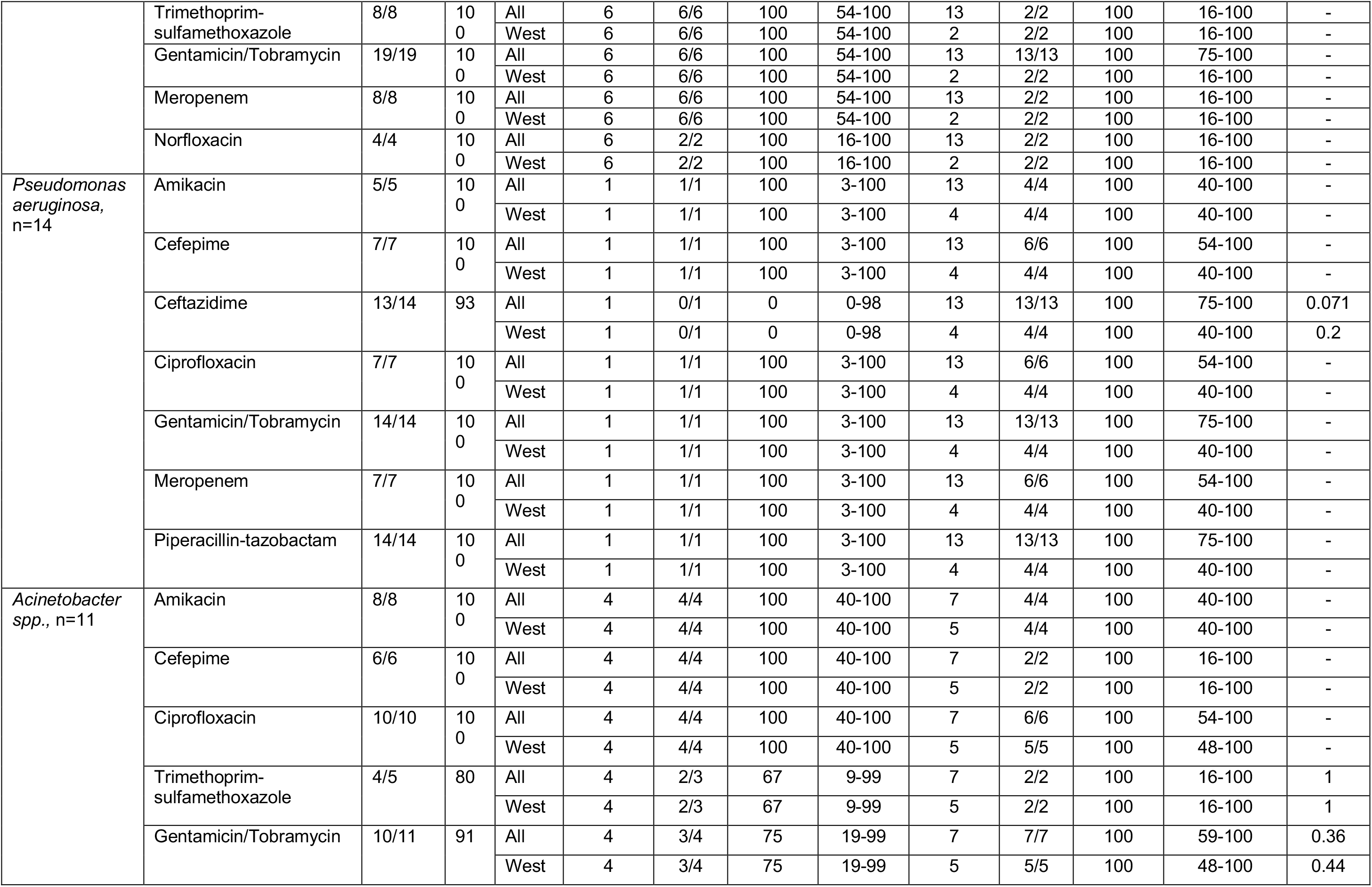
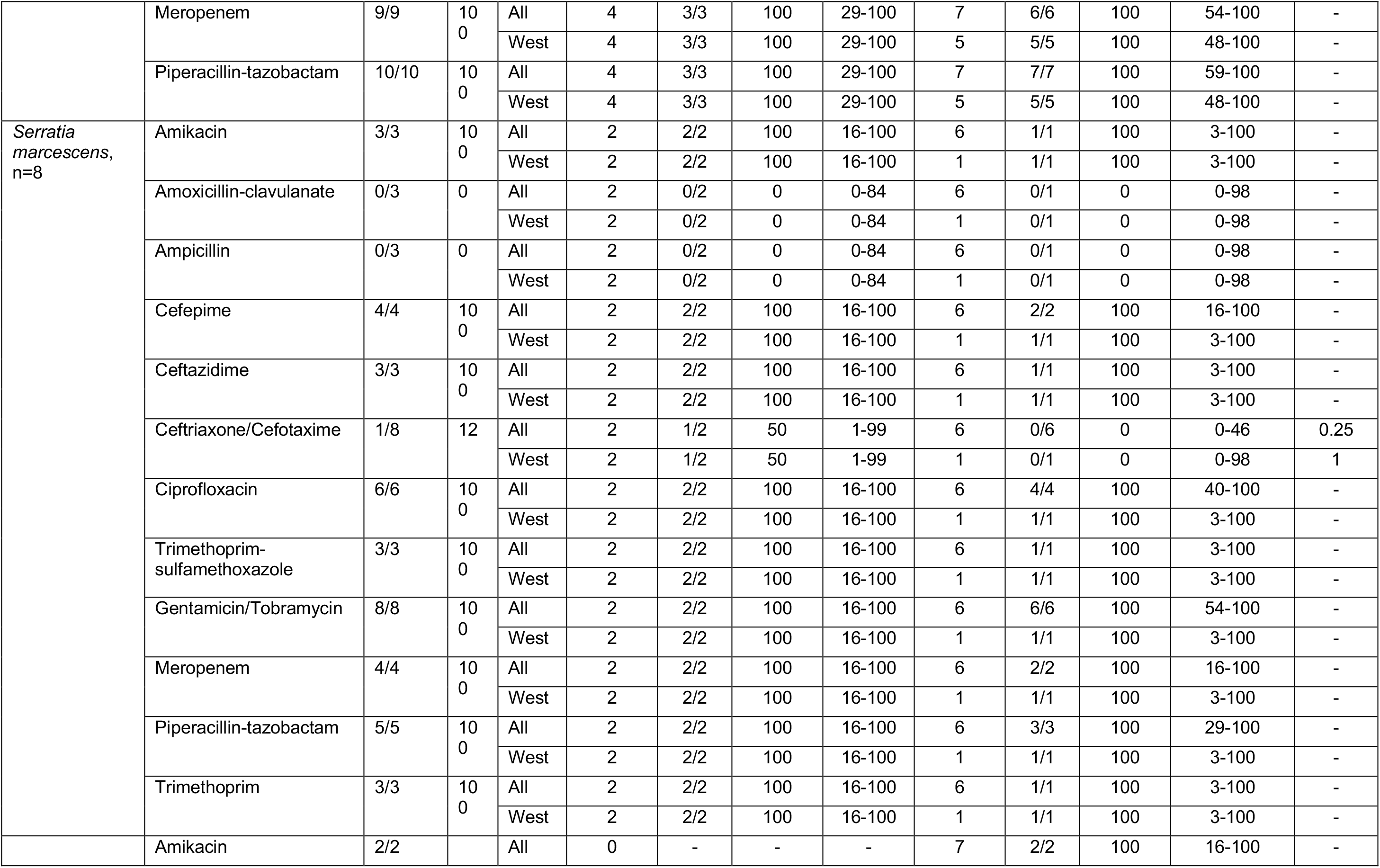
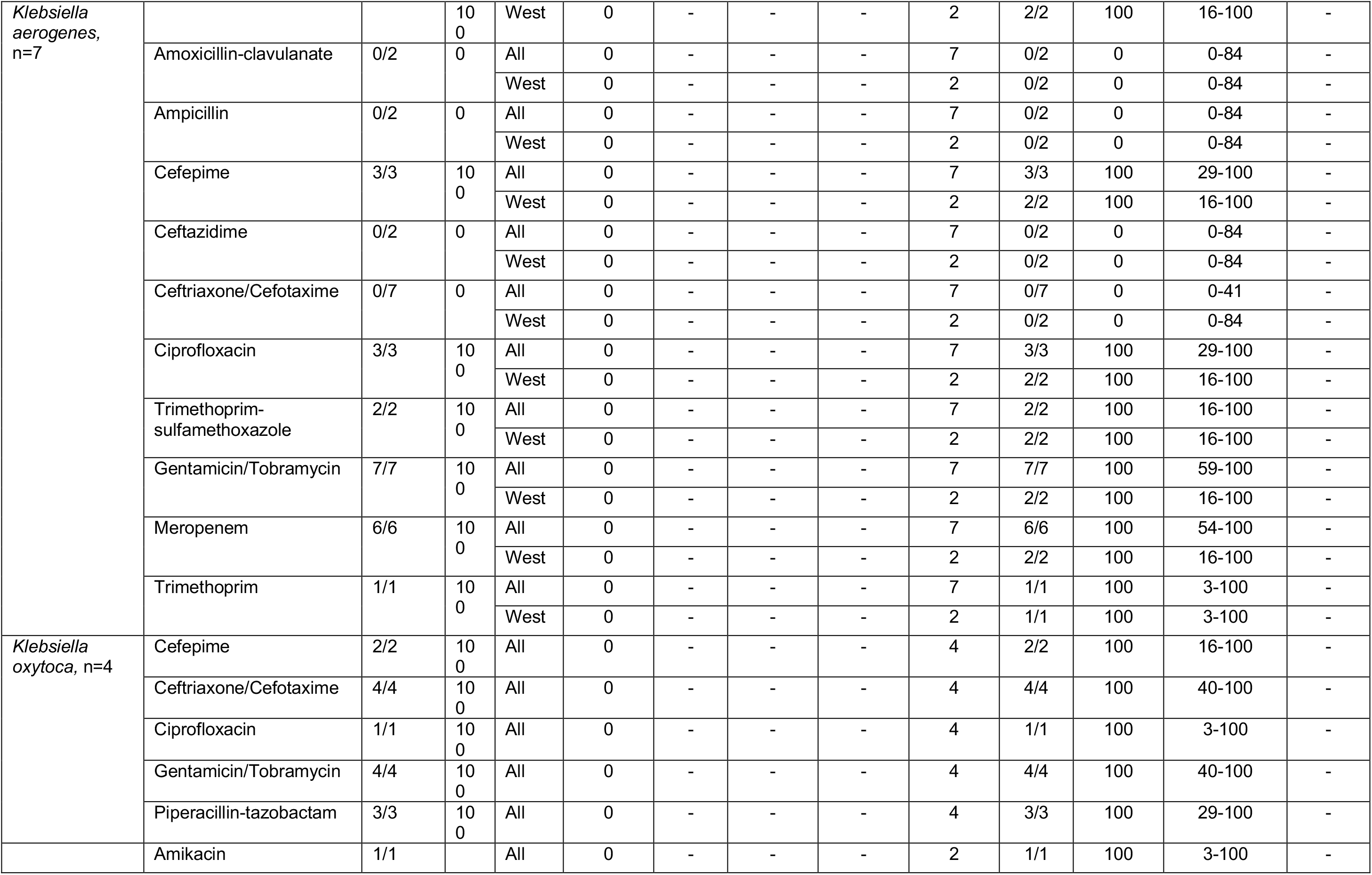
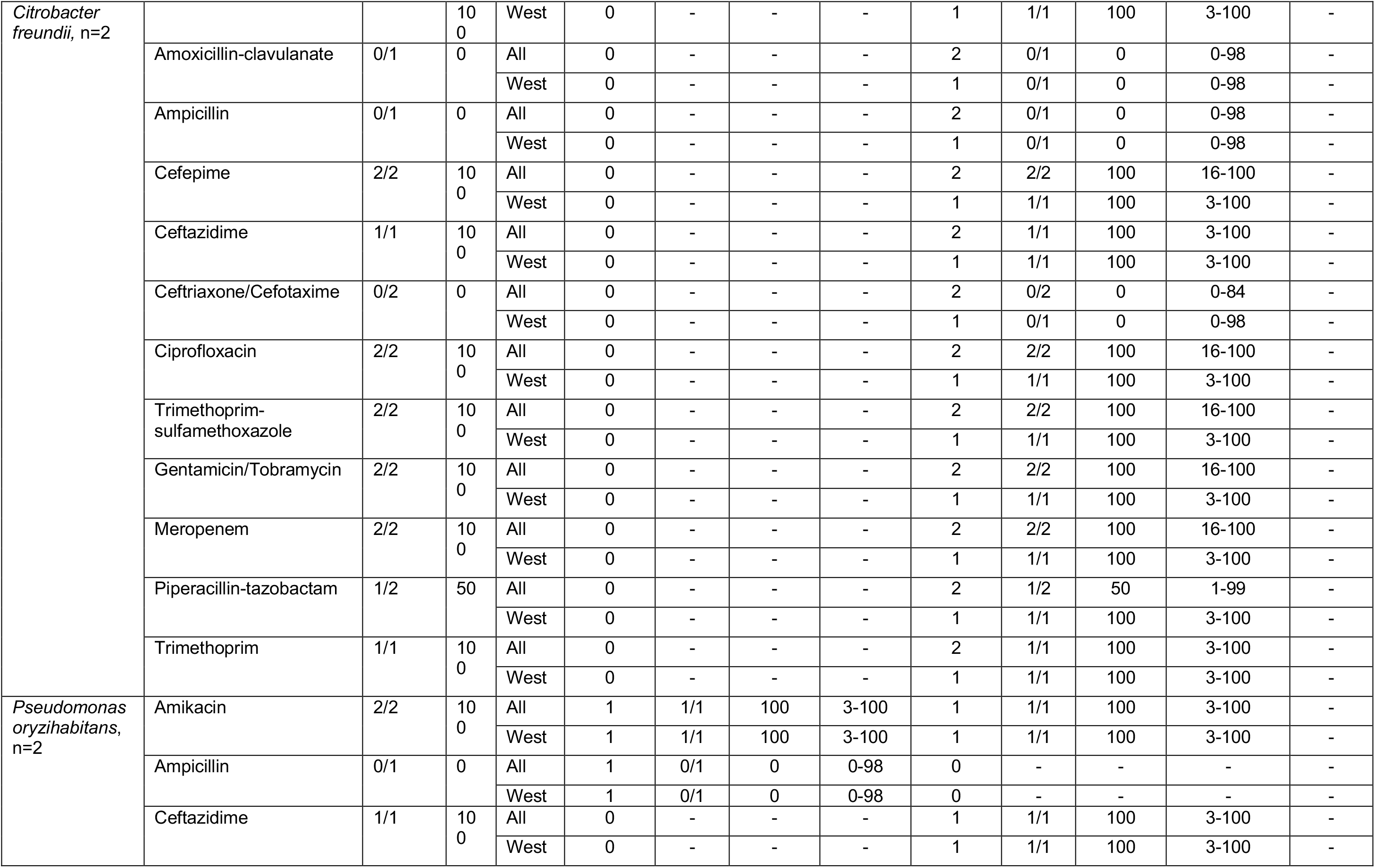
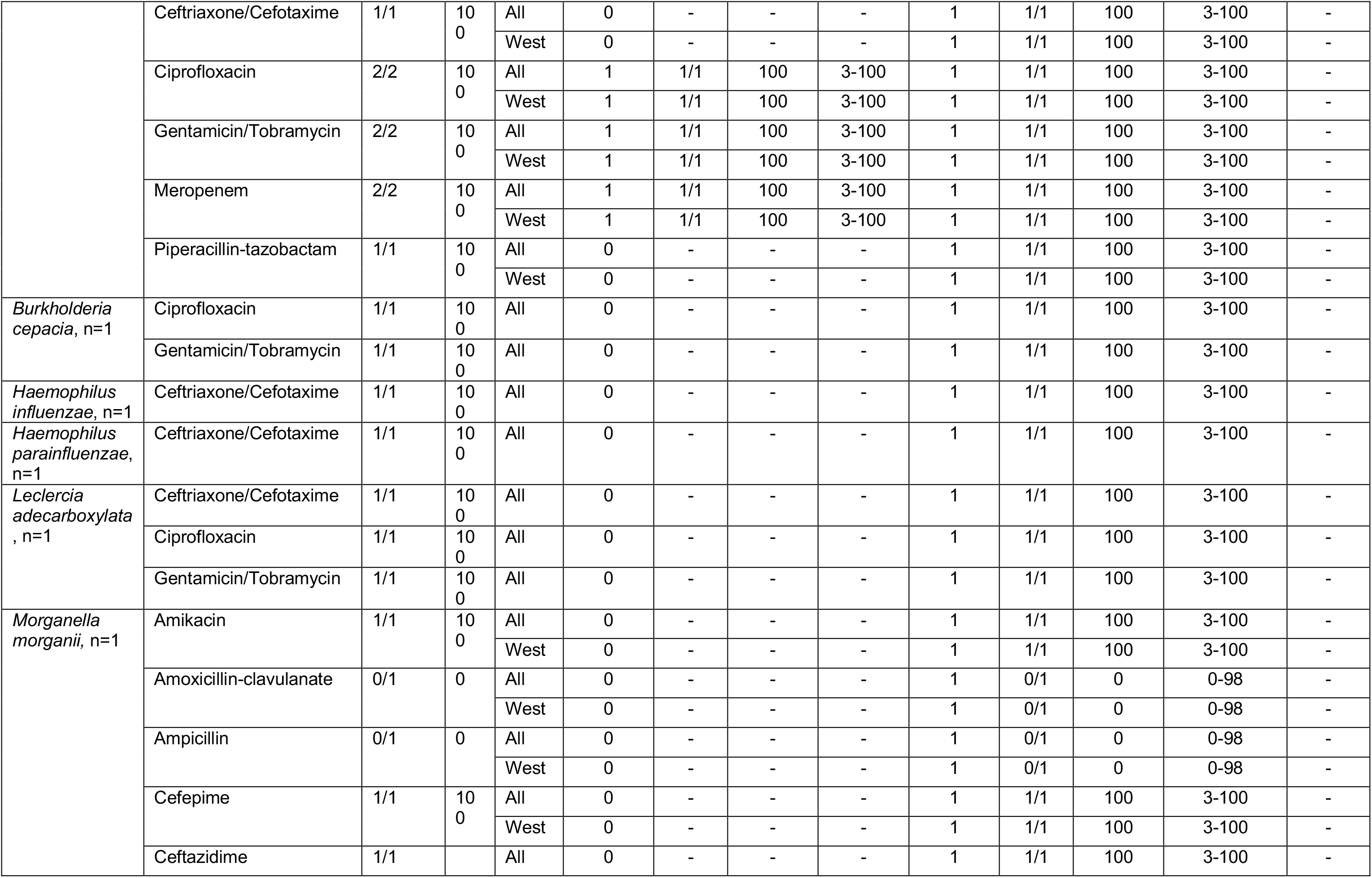
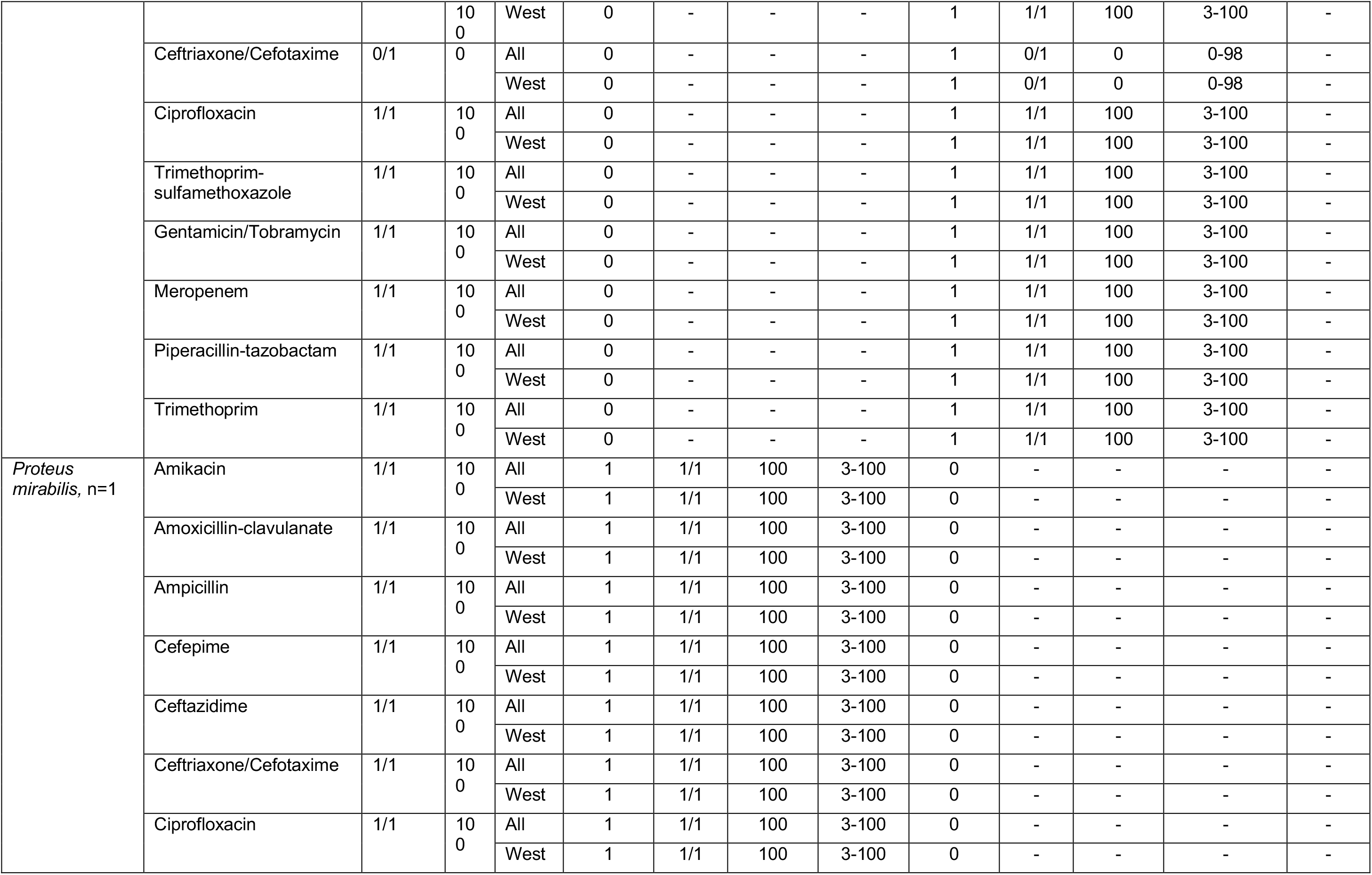
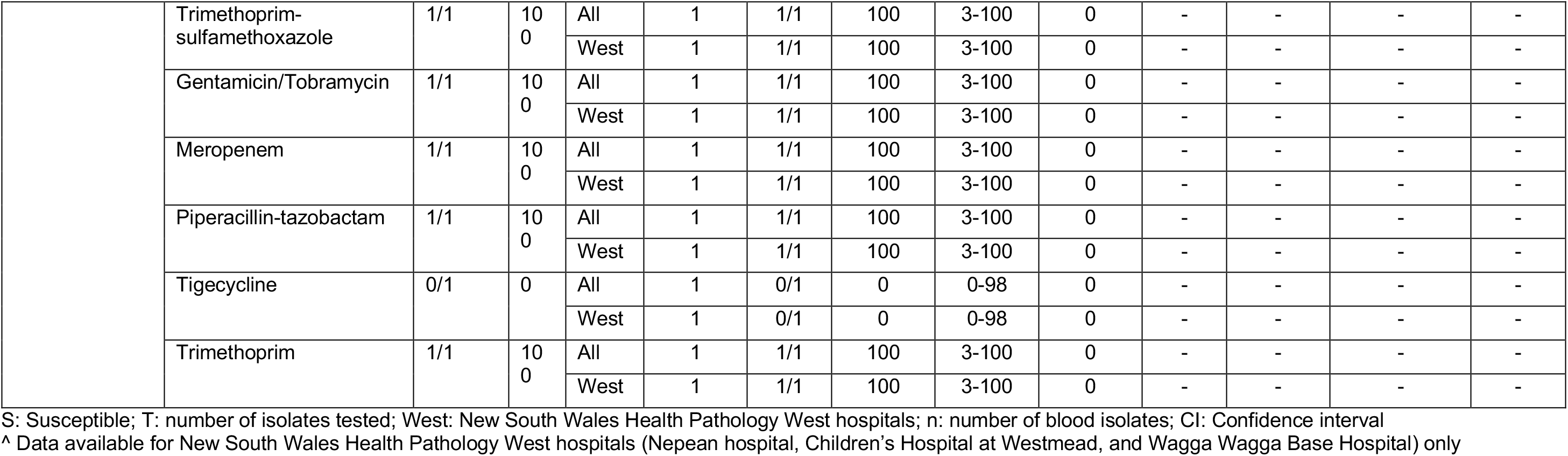
Gram-negative Blood Culture Antimicrobial Susceptibility Profiles.

**Supplementary Table 5.** Gram-negative Cerebrospinal Fluid Culture Antimicrobial Susceptibility Profiles.

| Pathogen | Antibiotic<br>(number susceptible/number tested; % susceptible) |  |  |  |  |  |  |  |  |  |  |
| --- | --- | --- | --- | --- | --- | --- | --- | --- | --- | --- | --- |
|  | Ampicillin | AMX-CLAV | Ceftriaxone | Cefepime | Ceftazidime | TMP-SMX | Ciprofloxacin | Gentamicin | Amikacin | PIP-TAZ | Meropenem |
| *Escherichia coli*, n=7 | 1/2 (50) | 2/2 (100) | 6/6 (100) | 2/2 (100) | 2/2 (100) | 2/3 (67) | 4/4 (100) | 6/6 (100) | 2/2 (100) |  | 3/3 (100) |
| *Enterobacter cloacae* complex n=2 |  |  | 0/2 (0) | 2/2 (100) |  |  |  | 2/2 (100) |  | 0/1 (0) |  |
| *Pseudomonas aeruginosa*, n=2 |  |  |  |  | 2/2 (100) |  | 1/1 (100) | 2/2 (100) |  | 1/1 (100) | 1/1 (100) |
| *Pseudomonas luteola*, n=1 |  |  |  |  | 1/1 (100) |  | 1/1 (100) | 1/1 (100) |  |  |  |
| *Klebsiella oxytoca*, n=1 |  |  | 1/1 (100) |  |  |  | 1/1 (100) | 1/1 (100) |  |  |  |
| *Neisseria meningitidis*, n=1 |  |  | 1/1 (100) |  |  |  |  |  |  |  |  |
| *Serratia liquefaciens*, n=1 |  |  | 1/1 (100) |  |  |  | 1/1 (100) | 1/1 (100) |  |  |  |
| *Serratia marcescens*, n=1 |  |  | 0/1 (0) |  |  |  | 1/1 (100) | 1/1 (100) |  |  |  |

AMX-CLAV: Amoxicillin-clavulanate; Ceftriaxone: Ceftriaxone or Cefotaxime; TMP-SMX: Trimethoprim-sulfamethoxazole; Gentamicin: Gentamicin or Tobramycin; PIP-TAZ: Piperacillin-tazobactam

**Supplementary Table 6.**
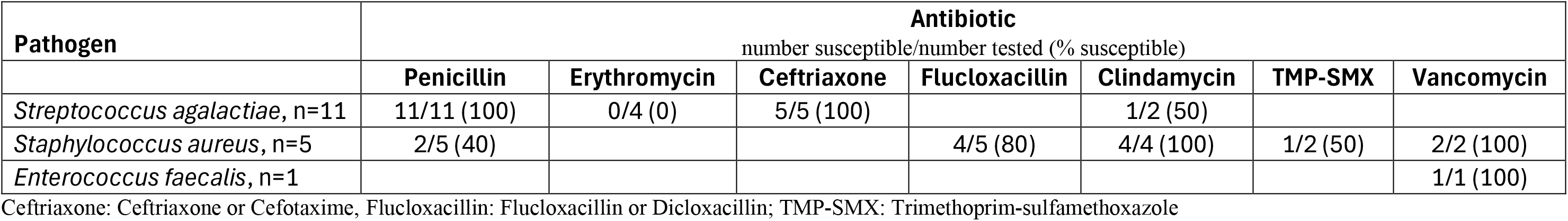
Gram-positive CSF isolate antimicrobial susceptibility profiles.

Gram-positive CSF isolate antimicrobial susceptibility profiles
| Pathogen | Antibiotic |  |  |  |  |  |  |
| --- | --- | --- | --- | --- | --- | --- | --- |
|  | number susceptible/number tested (% susceptible) |  |  |  |  |  |  |
|  | Penicillin | Erythromycin | Ceftriaxone | Flucloxacillin | Clindamycin | TMP-SMX | Vancomycin |
| *Streptococcus agalactiae*, n=11 | 11/11 (100) | 0/4 (0) | 5/5 (100) |  | 1/2 (50) |  |  |
| *Staphylococcus aureus*, n=5 | 2/5 (40) |  |  | 4/5 (80) | 4/4 (100) | 1/2 (50) | 2/2 (100) |
| *Enterococcus faecalis*, n=1 |  |  |  |  |  |  | 1/1 (100) |

Ceftriaxone: Ceftriaxone or Cefotaxime, Flucloxacillin: Flucloxacillin or Dicloxacillin; TMP-SMX: Trimethoprim-sulfamethoxazole

